# Cerebral Visual Impairment: genetic diagnoses and phenotypic associations

**DOI:** 10.1101/2023.10.02.23296432

**Authors:** Emogene Shaw, Genomics England Research Consortium, Ian Flitcroft, Richard Bowman, Kate Baker

**Author notes:** **Competing interests:** Authors have no competing interests to declare. **DATA AVAILABILITY STATEMENT:** No new data were generated by this research. See text for references to accessible data resources.

## Abstract

**Background:** Cerebral visual impairment (CVI) is the commonest form of paediatric visual impairment in developed countries. CVI can arise from a host of genetic or acquired causes, but there has been limited research to date on CVI in the context of genetic disorders.

**Methods:** We carried out a retrospective analysis of genotypic and phenotypic data for participants with CVI within the DECIPHER database and 100,000 Genomes Project (100KGP).

**Results:** 158 individuals with CVI were identified across both cohorts. Within this group, pathogenic or likely pathogenic sequence variants in 173 genes were identified. 25 of these genes already have known associations with CVI, whilst the remaining 148 are candidate genes for this phenotype. Gene Ontology analysis of the CVI gene sets from both DECIPHER and 100KGP suggest that CVI has a similar degree of genetic heterogeneity to other neurodevelopmental phenotypes, and a strong association with genetic variants converging on ion channels and receptor functions. Individuals with a monogenic disorder and CVI have a higher frequency of epilepsies and severe neurodisability than individuals with a monogenic disorder but not CVI.

**Conclusion:** This study supports the availability of genetic testing for individuals with CVI alongside other neurodevelopmental difficulties. It also supports the availability of ophthalmological screening for individuals with genetic diagnoses linked to CVI. Further studies could elaborate on the links between specific genetic disorders, visual maturation and broader neurodevelopmental characteristics.

**What is known about the subject?:**

- CVI is the most prevalent form of paediatric-onset visual impairment in the developed world.
- CVI commonly occurs alongside other neurodevelopmental phenotypes such as epilepsies, global developmental delay, cerebral palsy, ADHD and autism.
- CVI is known to have genetic aetiology, which is often undiagnosed.

**What this study adds?:**

- This study reports CVI-associated genetic diagnoses and phenotypic associations for two large cohorts
- Gene ontology annotations have been analysed for candidate CVI genes

**How this study might affect research, practice or policy:**

- This study has implications for genetic testing in children with CVI
- This study has implications for CVI screening in children with a genetic diagnosis

## INTRODUCTION

Cerebral visual impairment (CVI), also known as Cortical Visual Impairment, describes childhood-onset vision problems caused by damage to, or malfunctioning of, the cerebral components of the visual system. CVI can present with diverse symptoms including decreased visual acuity, visual field defects and visual cognitive dysfunctions arising from the dorsal and ventral processing streams [1–3]. CVI is the most prevalent form of paediatric-onset visual impairment in the developed world [1]. A recent UK cross-sectional population survey of 2298 children aged 5 to 11 years found that 3.4% had at least one CVI-related vision problem, indicating that CVI is more prevalent than previously thought [4]. CVI can be an isolated phenotype, but commonly occurs alongside other neurodevelopmental phenotypes such as epilepsies, global developmental delay, cerebral palsy, ADHD and autism spectrum disorders [5–7]. These co-occurring phenotypes emphasise the complex interplay between visual development and other neurological, cognitive and functional domains. Recognising CVI is important, whether isolated or part of a broader neurodevelopmental condition, because implementation of vision-appropriate environmental modifications and support strategies can promote cognitive development, educational attainment and social-emotional well-being [8 9].

CVI can arise due to numerous risk factors which compromise brain development during sensitive periods for visual development. Well-established acquired risk factors include prematurity, hydrocephalus, perinatal hypoxia and congenital infection [10 11]. However, primary genetic causes have received less attention, in terms of research or clinical evaluation. Two studies from the Netherlands by Bosch et al highlighted the contribution of genomic disorders to the aetiology of CVI. In a retrospective study, 7% of a well-characterised CVI cohort had at least one clinically significant chromosomal abnormality [12]. Building on this finding of a high genomic diagnostic yield, trio exome sequencing was carried out for 25 cases of idiopathic CVI, which identified a definite or potential genetic aetiology in 16 individuals [13]. Amongst specific findings were variants in 4 genes already reported to be associated with CVI (*AHDC1, NGLY1, NR2F1,* and *PGAP1*), and variants in 19 further candidate genes. This study reinforced that a significant proportion of individuals with CVI are likely to have an underlying genomic diagnosis, with extensive genomic heterogeneity.

Genetic diagnosis can provide clinically relevant insights into aspects of phenotypic heterogeneity within CVI. Genetic versus acquired CVI may be associated with a somewhat different profile of ophthalmological signs and visual dysfunction [14]. Identifying a genetic cause of CVI may have prognostic significance for the trajectory of visual function i.e. identification of a neurodegenerative condition or associated retinal pathology, which in turn may have important clinical management implications. Genetic diagnosis may also make a contribution to predicting whether a young child identified with visual difficulties is likely to experience problems in other neurodevelopmental domains or be at risk of co-morbidities such as epilepsy. In future, understanding the links between specific gene variants, visual development and other functional domains may lead to targeted intervention strategies to improve vision and associated outcomes. However, at present the evidence base to inform clinical genetic testing and post-diagnostic management for people with CVI of genetic origin is currently lacking.

In the current paper, we sought to build on previous studies by investigating genetic diagnoses associated with CVI, in two relevant largescale datasets - 100K Genomes Project (https://www.genomicsengland.co.uk/about-genomics-england/the-100000-genomes-project/) and DECIPHER [15]. Our primary objectives were to examine the catalogue of identified genomic variants for evidence of a consistent link with visual phenotypes, and determine via gene ontology enrichment analysis whether genes associated with CVI converge on a discrete set of biological processes, distinctive from neurodevelopmental disorders as a whole. In addition, we examined the co-occurring phenotypic characteristics of individuals with CVI of known genetic origin, to provide some initial guidance on which patients with CVI are most likely to have a monogenic diagnosis, and which patients with neurodevelopmental disorders are most likely to have CVI.

## MATERIALS AND METHODS

### Data sources for identification of people with CVI and genetic diagnoses

Individuals with CVI were identified within two large genomic datasets. The DECIPHER database is an international repository of clinically-diagnosed genomic variants, alongside clinician-reported Human Phenotype Ontology (HPO) terms (www.deciphergenomics.org). Open access data for all DECIPHER entries with single nucleotide variants (not copy number variants) was accessed on 21/2/2021 under a collaborative agreement between Wellcome Sanger Institute and University of Cambridge (K. Baker). To create a cohort of individuals with CVI and a likely genetic diagnosis, DECIPHER data was filtered to include individuals with CVI as a reported phenotype, and at least one Pathogenic or Likely Pathogenic variant. A similar process and comparable inclusion criteria were applied to identify a parallel cohort within the 100,000 Genomes Project (100KGP). 100KGP is a large UK-based research study which recruited individuals with rare disorders of unknown aetiology and applied whole genome sequencing and a consistent pipeline with clinically-relevant panel analyses, to diagnose known disorders and discover new causes of disease. Since its completion, 100KGP has generated a large research environment containing the genomic and phenotypic information of 33,029 individuals with rare disorder presentations [16]. Individuals with CVI in the 100KGP dataset were recruited under the rare disease category ‘neurology and neurodevelopmental disorders’. Individuals with CVI listed as a clinician-reported HPO term were identified by filtering the ‘rare_diseases_participant_phenotype’ data in LabKey. Genomic data were filtered within LabKey to include only Tier 1 and Tier 2 (corresponding to pathogenic and likely-pathogenic) variants, and merged with each participant’s complete list of HPO annotations. Data was analysed within the Genomics England Research Environment, utilising R scripts and Excel, and results were exported from the interface following review committee approval.

### Cataloguing of genetic diagnoses associated with CVI

The diagnostic yield of whole genome sequencing in people with CVI (in the context of broader neurodevelopmental or neurological presentations) was estimated within the 100KGP cohort as [number of participants with CVI and a Tier 1 or Tier 2 variant] divided by [number of participants with CVI]. To generate a catalogue of genetic diagnoses associated with CVI (Supplementary Tables 1 and 2), we collated a list of variants identified in people with CVI from the DECIPHER and 100KGP cohorts. To evaluate whether catalogued genes are already known to be associated with CVI, this list was cross-checked against the HPO website (accessed 14/04/2021). Using PubMed, literature on genes which appeared across both CVI cohorts was searched to establish any known connections between variants within the genes and CVI or other visual phenotypes.

### Gene ontology enrichment analysis

Given the genetic heterogeneity of CVI, and extreme genetic heterogeneity of neurodevelopmental disorders more broadly, we wanted to find out if there were any shared functional characteristics amongst genetic diagnoses in the DECIPHER and 100KGP CVI cohorts, which were distinctive from functional characteristics of developmental disorders in general. This would provide supportive evidence that CVI-associated genetic diagnoses converge on shared developmental mechanisms. ShinyGO v0.77 was used to conduct gene ontology enrichment analysis separately for the DECIPHER and 100KGP CVI gene lists. GO molecular function annotations were statistically tested to determine if they were over-represented within the gene sets via the hypergeometric test and Benjamin Hochberg FDR correction. The p-value cut off was set to 0.05 and only the top 30 pathways selected by FDR were visualised, to aid interpretation of the most significant results.

To compare CVI-associated GO enrichment networks to GO enrichment networks associated with developmental disorders more broadly, control cohorts were created from both datasets. Within the DECIPHER dataset this was done in Excel via the column filter tools to remove those with CVI, and to only include those with a ‘Pathogenic’ or ‘Likely Pathogenic’ variant in a gene not within the candidate CVI gene list. Within 100KGP, this was done by filtering the ‘rare_diseases_participant_phenotype’ data to exclude those with CVI listed, and merging this with the ‘tiering_data’ for individuals recruited under ‘neurodevelopmental disorders’ with variants rated as Tier 1 or Tier 2 in a gene not within the candidate CVI gene list. The INDEX and RAND functions in excel were utilised to randomly select and create 10 control gene lists with the same number of genes as the CVI cohort gene lists. These control gene lists were then analysed for enriched pathways in ShinyGO using the same settings.

### Evaluation of phenotypes co-occurring with CVI of genetic origin

To highlight clinical features that are most likely to co-occur amongst individuals with CVI of genetic origin and which differentiate CVI-associated diagnoses from other neurogenetic diagnoses, we analysed HPO terms in addition to CVI, for the cohorts within both DECIPHER and 100KGP. We compared the frequency of the top 20 HPO terms within each CVI cohort to control cohorts. The DECIPHER control cohort was created by selecting individuals with a documented pathogenic or likely pathogenic variant in a gene not within the candidate CVI gene list and who did not have CVI reported as a phenotype. The 100KGP control cohort was created by selecting individuals who were recruited under rare disease ‘neurology and neurodevelopmental disorders’ and had a tier 1 or tier 2 variant in a gene not within the candidate CVI gene list and no CVI reported. Fisher’s exact test with Holm-Bonferroni correction for multiple comparisons were used for statistical analysis.

## RESULTS

### CVI-associated genetic variants in DECIPHER

Within DECIPHER, 61 individuals were reported to have CVI. 46 sequence variants had been classified as pathogenic or likely pathogenic variants by reporting clinical laboratories, in 42 individuals. This produced a list of 36 candidate genes for CVI, listed in Supplementary Table 1. Pathogenic or likely pathogenic variants in 5 genes (*GRIN2B, IQSEC2, SMC1A, GRIN1* and *ITPR1)* were found in more than 1 individual. Single instances of pathogenic variants were found in 16 other genes, and likely pathogenic variants in 15 other genes. Of these 36 genes, 12 are already known to be associated with CVI, according to HPO (18).

### CVI-associated genetic variants in GEL

Within the 100KGP rare disease cohort (neurodevelopmental or neurological referral criteria), 97 individuals were reported to have CVI. 71 of these individuals had at least one Tier 1 or Tier 2 genetic variant. However, it should be noted that a very large number of gene variants were identified (30 Tier 1 and 211 Tier 2 variants across 144 genes within the 71 individuals i.e. average of 3 potentially pathogenic variants per individual) and not all variants will be pathogenic after further molecular and clinical evaluation. Amongst these 144 genes, 17 are known to be associated with CVI, according to HPO [17]. Tier 1 or Tier 2 variants in 7 genes (*PITRM1, KCNT1, CACNA1A, SHANK3, KANSL1, GRIN2B* and *WDR73)* were found in more than 1 individual. Supplementary Table 2 contains the full list of genes and count of individuals with variants in said genes.

### Overlap of genetic diagnoses within Decipher and GEL

Seven genes were identified in both the DECIPHER and 100KGP cohorts, lending additional support to their association with CVI (*GRIN2B, TCF4, KAT6A, PDHA1, KIF1A, FOXG1* and *RARS2)*. Individual variants were checked to ensure independence of the two cohorts – no specific variant appeared in both 100KGP and DECIPHER. Of these seven genes, three (*PDHA1, FOXG1, RARS2*) are not currently associated with CVI or visual impairment according to HPO. Review of published case series for these seven diagnoses (Supplementary Table 3) highlighted that three genes (*FOXG1, GRIN2B, KIF1A*) have previously been associated with CVI in between 7% and 41% of reported cases; two genes (*RARS2, KAT6A*) have been associated with visual impairment in 19% and 65% of patients respectively, but CVI was not specified by authors; ophthalmological phenotypes (strabismus, refractive errors) are reported in a high proportion of patients with *TCF4* (Pitt Hopkins Syndrome) and *PDHA1* (mitochondrial disease) variants, but case series have not commented on functional vision. Of note, four genes (*FOXG1, RARS2, KAT6A, KIF1A*) have reported associations with a combination of ocular, ophthalmological and functional visual phenotypes, indicating complex ophthalmological assessment and management needs.

### Gene Ontology analysis

GO analysis of candidate gene lists collected from both cohorts demonstrated networks of significantly enriched molecular functions (meaning that related gene functional annotations were represented above expectation for size of gene set). The DECIPHER CVI gene list generated one network (Figure 1a), corresponding to neuronal ion channel and receptor functions. Smaller networks of enriched function were identified for four out of the ten control gene sets (Supplementary Figure 1), none of which involved the same enriched functions as CVI. This indicates a higher functional homogeneity within the CVI gene set than control gene sets, noting that DECIPHER gene sets were created across all developmental disorder types not selected for neurodevelopmental phenotypes. The most significantly enriched molecular function within the DECIPHER gene list was glutamate-gated calcium ion channel activity, with a fold enrichment value of 414.5, and this term was not found to be enriched in any of the 10 control gene lists.

**Figure 1:**
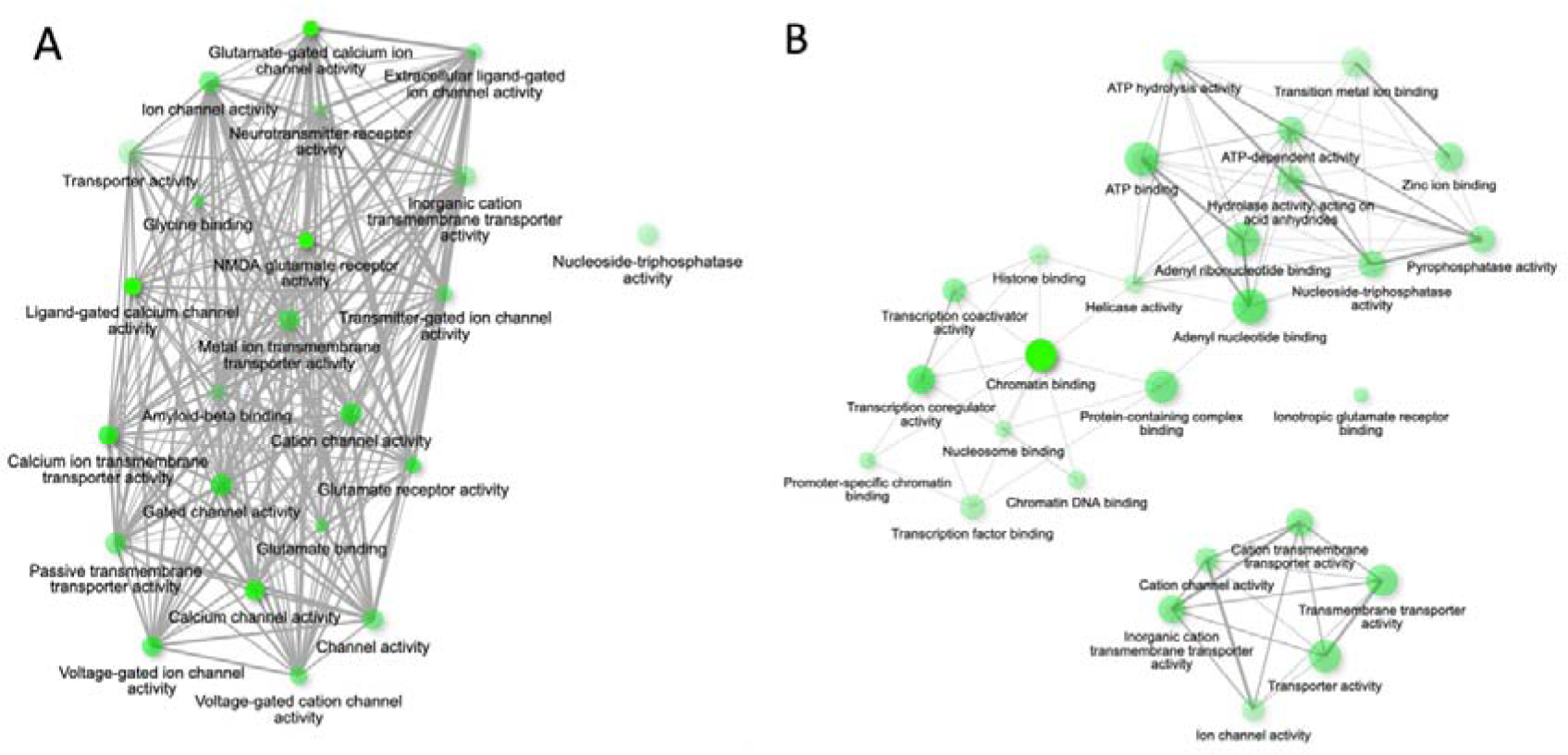
ShinyGO networks produced for CVI candidate gene sets. A=DECIPHER data, B=100KGP data. Nodes represent GO molecular function annotations and transparency of nodes relate to significance (with more opaque nodes meaning most significant) and are joined by edges if they share 20% of the same genes (thicker edges relate to more shared genes)

The CVI gene list produced from 100KGP highlighted two networks of over-represented molecular functions corresponding to chromatin binding and transcriptional regulation; and ion channel or transmembrane transporter activity (Figure 1b). In contrast to the DECIPHER analysis, enriched networks were also identified within all control groups, which were selected for neurodevelopmental recruitment criteria (Supplementary Figure 2). Chromatin binding functions were significantly enriched in 6 of the 10 control gene sets, and ion channel or transporter-related functions were enriched in 9 out of 10 control genes sets. The single most enriched molecular function term within the CVI gene list (not forming a network with other pathway terms) was Ionotropic glutamate receptor binding (23 fold enrichment, based on 4 genes within the set). This term was enriched in only 1 of the 10 control gene lists. In summary, gene functional heterogeneity was found to be similar overall between 100KGP participants recruited for neurodevelopmental presentations with and without CVI, with a similar representation of genes related to chromatin regulation and ion channel-related functions within the CVI and control groups. However, based on observations across both 100KGP and DECIPHER, there is evidence in support of an association between glutamatergic receptor-related functions and CVI.

### Phenotype analysis across cohorts

Figure 2a and Supplementary Table 4 display the 20 HPO terms most frequently reported for the DECIPHER CVI cohort (individuals with CVI and a pathogenic or likely pathogenic variant), in comparison to their reported frequency within the DECIPHER control cohort (individuals without CVI, with a pathogenic or likely pathogenic variant). The HPO terms “seizure”, “epileptic spasm” and “gastrostomy tube feeding in infancy” were all reported at significantly higher frequency for CVI participants. Although frequency of “global developmental delay” and “intellectual disability” did not significantly differ between groups, the CVI group were more likely to be reported as having severe or profound developmental delay (29% and 12% of the CVI group, respectively).

**Figure 2:**
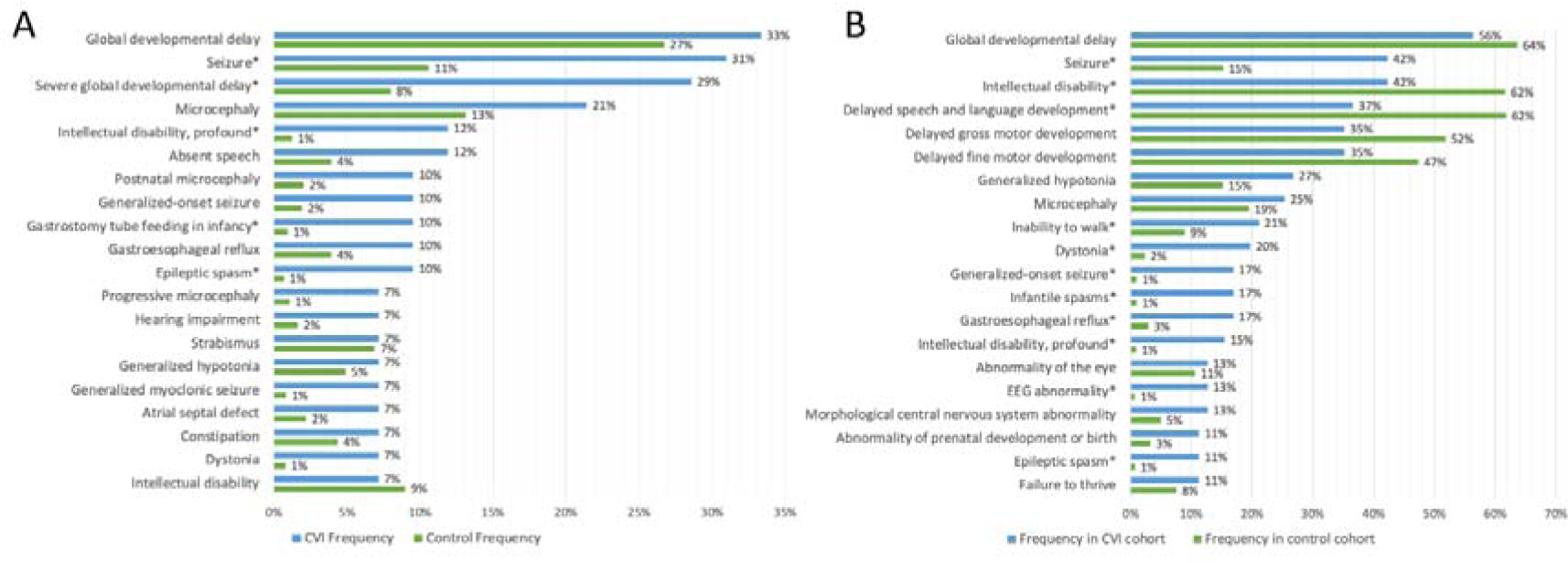
Top 20 HPO term frequencies in CVI and control groups. A=DECIPHER data, B=100KGP data. Significant differences identified with an asterisk.

Figure 2b and Supplementary Table 5 shows the 20 HPO terms most frequently reported for the CVI cohort within 100KGP (participants with CVI and a Tier 1 or 2 variant), and their prevalence in comparison to the 100KGP control cohort (participants recruited for a neurodevelopmental indication, without CVI, and a Tier 1 or 2 variant). CVI participants were more like to have seizures (of any type), with a particularly high proportion reported to have infantile spasms (17% in the CVI group versus 1% in the control group). There were no significant differences between groups in the likelihood of a CNS morphological abnormality or microcephaly, but higher likelihood of EEG abnormality within the CVI group. Dystonia (20% versus 2%) and gastro-oesophageal reflux (17% versus 3%) were also significantly different between groups. Regarding neurodevelopmental characteristics, participants with CVI were less likely to be annotated with the HPO term “intellectual disability”, but more likely to have the term “intellectual disability-profound”, and more likely to have the term “inability to walk”. Interestingly, there was a significantly higher proportion of individuals within the 100KGP CVI versus non-CVI group reported to have abnormality of prenatal development or birth.

## DISCUSSION

In this paper, we review the genetic diagnoses and phenotypic characteristics of individuals with CVI in the context of monogenic conditions. Improved access to genomic diagnostics will lead to a rapid increase in the size of the population of individuals with CVI of known genetic cause, and reduction in age at receiving a genetic diagnosis. This leads to new opportunities for advancement in our understanding of developmental visual impairments and improvements in clinical care. Hence, the data presented here provide early insights into this population, to be expanded and enhanced in further studies.

Our first objective was to examine the catalogue of identified genomic variants in individuals reported to have CVI across two cohorts – one ascertained after genetic diagnosis (DECIPHER) and one ascertained prior to diagnosis (100KGP). This search identified 173 genes harbouring likely pathogenic or pathogenic variants, in a total of 132 individuals reported to have CVI. This suggests a potentially high diagnostic yield for individuals presenting with CVI alongside other developmental phenotypes. Of these 173 genes, 25 are already recognised to be associated with CVI according to HPO. 38 genes were implicated in more than one case across cohorts; 135 genes had only one instance of a pathogenic or likely pathogenic variant in either cohort. Only 7 genes appeared in both the 100KGP and DECIPHER cohorts, and review of literature indicates that there has been variable reporting of CVI in previous case series of these 7 conditions. CVI may have been over-shadowed by a focus on peripheral ocular phenotypes in some conditions, and focus on autism as an explanation for poor eye contact in others. CVI has only been investigated in depth in one of these conditions (*FOXG1*) where it is highly prevalent [18] and associated with abnormal visual evoked potentials [19], which could be a useful methodology for investigation of other conditions identified in the current study.

As expected, these results confirm the very high degree of genetic heterogeneity of complex neurogenetic conditions which involve CVI. These observations emphasise the difficulty of predicting a causative diagnosis during pre-test counselling, and difficulty of predicting phenotypic presentations in ultra-rare conditions, which may have highly individualised effects depending on each individual’s specific variant and their developmental course. A limitation of this aspect of our study is that further evaluation of the pathogenicity of this large catalogue of variants was beyond scope – this would require further clinical evaluation to obtain fine-grained and longitudinal phenotypic information, confirmatory biochemical or other clinical investigations, and functional studies for novel variants. The presence of multiple potential causative variants within individuals is an increasingly common clinical scenario, where linking each variant to specific phenotypes is very challenging and a conjoint or burden effect contributing to complex developmental presentation may be more realistic.

In view of the high degree of genetic heterogeneity associated with CVI, we wanted to establish whether there is any evidence for convergence of CVI-associated genetic diagnoses on molecular functional pathways. Such convergence might assist in interpreting the pathogenicity of novel CVI-associated variants, identifying additional CVI-associated candidate genes, and highlighting potential pathophysiological pathways relevant to CVI. Gene ontology analysis of the diagnostic catalogues harvested from both cohorts identified networks of over-represented functions. In comparison to GO analysis of 10 randomly selected gene lists from DECIPHER, there was evidence for higher functional homogeneity within the CVI-associated gene list, because there were two strongly inter-connected and enriched networks within the CVI gene list, but much less extensive enrichment and networking within the control gene lists. However, this distinction did not arise in analysis of the 100KGP CVI and control gene lists, where control gene lists were drawn from participants recruited for neurological and neurodevelopmental indications. This suggests that the genetic heterogeneity within individuals with CVI is lower than observed across non-CNS or multi-system developmental disorders (DECIPHER) but no different from heterogeneity across CNS-related disorders. Comparing the specifically enriched functional annotations across both cohorts, it is apparent that genes involved in ion channel function and glutamatergic receptors (either variants in channel and receptor subunits, or variants in regulators of their expression and function) are functional “hotspots” for CVI. Further work is required to expand on this finding by investigating whether this relationship is mediated via seizures or via direct involvement of these protein complexes and neuronal functions in visual development. Other networks, notably those involved in chromatin organisation and transcriptional regulation, were enriched within both the CVI group and the neurodevelopmental control groups in GEL. In these conditions, a different set of mechanisms may contribute to visual development in particular structural brain development. For some conditions (*GRIN2B* in particular), both structural abnormalities of the cortex and electrophysiological abnormalities may contribute to CVI [20]. It is important to consider the evolving nature of the GO database when analysing functional annotations – network analyses will change and improve with further investigation of genes within the CVI-associated set.

Building on the observed genetic heterogeneity and functional network associations amongst individuals with CVI, we wanted to find out whether there was phenotypic convergence within this group, in comparison to individuals with monogenic developmental disorders not involving CVI. Summarising case-control analyses across both DECIPHER and 100KGP, individuals with CVI in the context of a monogenic disorder are more likely to have severe neurodisability when compared to individuals with monogenic developmental disorders in the absence of CVI. Early diagnosis of CVI in a child with a neurogenetic disorder may therefore be an indicator of cautiously poor neurodevelopmental prognosis, highlighting the need for more extensive and long-term educational and family support. There is also a strong association between CVI and seizures (particularly infantile spasms). This association may point toward pathophysiological mechanisms contributing to CVI. In conditions with high prevalence of seizures and CVI, disturbance to visual development may be a secondary consequence of seizures, or may arise due to shared molecular and electrophysiological mechanisms influencing visual maturation. A future study of the emergence of visual functions and visual impairments in children with monogenic epilepsies could provide significant insights into this question. In conditions with high prevalence of dystonia and CVI, disturbance to visual development may arise because of shared neuroanatomical substrate (subcortical-cortical systems affecting motor control and sensorimotor integration) as has been observed previously in cerebral palsy, potentially benefiting from a different mode of intervention.

Two broad clinical implications can be drawn from this study. Firstly, genetic testing should be offered to all individuals with CVI in the context of severe developmental delay, especially in the presence of epilepsy. A high yield of genetic diagnoses can be expected for this group. Such individuals already meet eligibility criteria for genetic testing in the UK, based on their neurodevelopmental characteristics (https://www.england.nhs.uk/publication/national-genomic-test-directories/). The offer of testing should not be restricted by an assumption of an acquired cause for CVI, even in the presence of another known risk factor such as prematurity. Genetic and acquired causes may well co-exist, and compound each other. We are not able to comment on the potential diagnostic yield and utility of offering genetic testing to individuals with CVI in the absence of co-occurring neurological or neurodevelopmental difficulties, since these individuals were not represented within either the DECIPHER or 100KGP cohorts. This could be the focus of a future genetic screening study.

A second clinical implication is that there should be a low threshold for CVI screening as a post-diagnostic assessment for individuals diagnosed with neurogenetic disorders, especially disorders which we have identified in multiple individuals with CVI across the examined genomic cohorts. Identification of visual needs, especially at a very early age, will lead to changes in family support and educational environment, likely to have positive impacts on long-term adaptive development, mental health and social inclusion. A systematic post-diagnostic study of CVI across these disorders is required to obtain information about prevalence and types of visual impairments and their progression with age and developmental progress.

Finally, the strong triangulation between channelopathies, early-onset seizure disorders and CVI should be a focus of translational research – there are active trials of novel therapeutics in these conditions, and the impact of interventions on visual function could be an important and useful outcome measure. Speculatively, understanding the pathophysiological basis of CVI in these conditions, and discovery of therapeutic interventions which can improve visual development, could be relevant to the larger population of individuals with CVI of heterogeneous genetic or acquired origin.

## Data Availability

No new data were generated by this research. See text for references to accessible data resources.

https://www.genomicsengland.co.uk/about-genomics-england/the-100000-genomes-project/

https://www.deciphergenomics.org/

## ETHICS STATEMENTS

### Patient consent for publication

Not required.

### Ethics approval

This study involves data previously collected for human participants, which was approved for Genomics England: HRA Committee East of England–Cambridge South (REC ref: 14/EE/1112). Participants gave informed consent to participate in the study before taking part.

## ACKNOWLEDGEMENTS

This research was made possible through access to the data and findings generated by the 100,000 Genomes Project. The 100,000 Genomes Project is managed by Genomics England Limited (a wholly owned company of the Department of Health and Social Care). The 100,000 Genomes Project is funded by the National Institute for Health Research and NHS England. The Wellcome Trust, Cancer Research UK and the Medical Research Council have also funded research infrastructure. The 100,000 Genomes Project uses data provided by patients and collected by the National Health Service as part of their care and support. This study makes use of data generated by the DECIPHER community. A full list of centres who contributed to the generation of the data is available from https://deciphergenomics.org/about/stats and via email from. DECIPHER is hosted by EMBL-EBI and funding for the DECIPHER project was provided by the Wellcome Trust [grant number WT223718/Z/21/Z]. Those who carried out the original analysis and collection of the data bear no responsibility for the further analysis or interpretation of the data. We are grateful to Dr Timothy Hearn, for advice on data management and analysis.

## REFERENCES

1. Fazzi E, Signorini SG, Bova SM, et al. Spectrum of visual disorders in children with cerebral visual impairment. J Child Neurol 2007;22(3):294–301 doi: 10.1177/08830738070220030801[published Online First: Epub Date]|.

2. van Genderen M, Dekker M, Pilon F, et al. Diagnosing cerebral visual impairment in children with good visual acuity. Strabismus 2012;20(2):78–83 doi: 10.3109/09273972.2012.680232[published Online First: Epub Date]|.

3. Lueck AH, Dutton GN, Chokron S. Profiling Children With Cerebral Visual Impairment Using Multiple Methods of Assessment to Aid in Differential Diagnosis. Semin Pediatr Neurol 2019;31:5–14 doi: 10.1016/j.spen.2019.05.003[published Online First: Epub Date]|.

4. Williams C, Pease A, Warnes P, et al. Cerebral visual impairment-related vision problems in primary school children: a cross-sectional survey. Dev Med Child Neurol 2021;63(6):683–89 doi: 10.1111/dmcn.14819[published Online First: Epub Date]|.

5. Ortibus E, Fazzi E, Dale N. Cerebral Visual Impairment and Clinical Assessment: The European Perspective. Semin Pediatr Neurol 2019;31:15–24 doi: 10.1016/j.spen.2019.05.004[published Online First: Epub Date]|.

6. Chokron S, Kovarski K, Zalla T, et al. The inter-relationships between cerebral visual impairment, autism and intellectual disability. Neurosci Biobehav Rev 2020;114:201–10 doi: 10.1016/j.neubiorev.2020.04.008[published Online First: Epub Date]|.

7. Chokron S, Dutton GN. Impact of Cerebral Visual Impairments on Motor Skills: Implications for Developmental Coordination Disorders. Front Psychol 2016;7:1471 doi: 10.3389/fpsyg.2016.01471[published Online First: Epub Date]|.

8. Dale N. Cerebral visual impairment-related vision problems in the classroom. Dev Med Child Neurol 2021;63(6):632 doi: 10.1111/dmcn.14837[published Online First: Epub Date]|.

9. Goodenough T, Pease A, Williams C. Bridging the Gap: Parent and Child Perspectives of Living With Cerebral Visual Impairments. Front Hum Neurosci 2021;15:689683 doi: 10.3389/fnhum.2021.689683[published Online First: Epub Date]|.

10. Pehere N, Chougule P, Dutton GN. Cerebral visual impairment in children: Causes and associated ophthalmological problems. Indian J Ophthalmol 2018;66(6):812–15 doi: 10.4103/ijo.IJO_1274_17[published Online First: Epub Date]|.

11. Handa S, Saffari SE, Borchert M. Factors Associated With Lack of Vision Improvement in Children With Cortical Visual Impairment. J Neuroophthalmol 2018;38(4):429–33 doi: 10.1097/WNO.0000000000000610[published Online First: Epub Date]|.

12. Bosch DG, Boonstra FN, Reijnders MR, et al. Chromosomal aberrations in cerebral visual impairment. Eur J Paediatr Neurol 2014;18(6):677–84 doi: 10.1016/j.ejpn.2014.05.002[published Online First: Epub Date]|.

13. Bosch DG, Boonstra FN, de Leeuw N, et al. Novel genetic causes for cerebral visual impairment. Eur J Hum Genet 2016;24(5):660–5 doi: 10.1038/ejhg.2015.186[published Online First: Epub Date]|.

14. Bosch DG, Boonstra FN, Willemsen MA, et al. Low vision due to cerebral visual impairment: differentiating between acquired and genetic causes. BMC Ophthalmol 2014;14:59 doi: 10.1186/1471-2415-14-59[published Online First: Epub Date]|.

15. Firth HV, Richards SM, Bevan AP, et al. DECIPHER: Database of Chromosomal Imbalance and Phenotype in Humans Using Ensembl Resources. Am J Hum Genet 2009;84(4):524–33 doi: 10.1016/j.ajhg.2009.03.010[published Online First: Epub Date]|.

16. Caulfield MD, Jim; Dennys, Martin; Elbahy, Leila; Fowler, Tom; Hill, Sue; et al. The National Genomics Research and Healthcare Knowledgebase., 2017.

17. Kohler S, Gargano M, Matentzoglu N, et al. The Human Phenotype Ontology in 2021. Nucleic Acids Res 2021;49(D1):D1207–D17 doi: 10.1093/nar/gkaa1043[published Online First: Epub Date]|.

18. Boggio EM, Pancrazi L, Gennaro M, et al. Visual impairment in FOXG1-mutated individuals and mice. Neuroscience 2016;324:496–508 doi: 10.1016/j.neuroscience.2016.03.027[published Online First: Epub Date]|.

19. Saby JN, Peters SU, Benke TA, et al. Comparison of evoked potentials across four related developmental encephalopathies. J Neurodev Disord 2023;15(1):10 doi: 10.1186/s11689-023-09479-9[published Online First: Epub Date]|.

20. Platzer K, Yuan H, Schutz H, et al. GRIN2B encephalopathy: novel findings on phenotype, variant clustering, functional consequences and treatment aspects. J Med Genet 2017;54(7):460–70 doi: 10.1136/jmedgenet-2016-104509[published Online First: Epub Date]|.

