## Supplementary Material for "Cerebral Visual Impairment: genetic diagnoses and phenotypic associations"

**Supplementary Table 1: CVI-associated genetic diagnoses in DECIPHER**

Key

* Genes already known to be associated with CVI on HPO website (https://hpo.jax.org/app/).

\# Genes represented in both the DECIPHER and 100KGP cohorts

| **Gene** | **Number of participants with CVI with a pathogenic variant reported** | **Number of participants with CVI with a likely pathogenic variant reported** |
| --- | --- | --- |
| GRIN2B*# | 2 | 1 |
| IQSEC2 | 1 | 1 |
| SMC1A* | 1 | 1 |
| GRIN1* | 0 | 2 |
| ITPR1 | 0 | 2 |
| ARV1 | 1 | 0 |
| CACNA1E* | 1 | 0 |
| CASK | 1 | 0 |
| CDKL5* | 1 | 0 |
| CEP290 | 1 | 0 |
| CERT1* | 1 | 0 |
| DNM1 | 1 | 0 |
| FOXG1 *# | 1 | 0 |
| GFM1 | 1 | 0 |
| GNAO1 | 1 | 0 |
| KCNQ2 | 1 | 0 |
| KDM5C | 1 | 0 |
| OPA1 | 1 | 0 |
| PLP1 | 1 | 0 |
| TCF4*# | 1 | 0 |
| TUBA1A | 1 | 0 |
| ATRX | 0 | 1 |
| DOCK6 | 0 | 1 |
| DYNC1H1* | 0 | 1 |
| FARS2 | 0 | 1 |
| GRIN2A | 0 | 1 |
| KAT6A*# | 0 | 1 |
| KIF1A*# | 0 | 1 |
| NAA10 | 0 | 1 |
| NGLY1 | 0 | 1 |
| PAX6 | 0 | 1 |
| PDHA1 # | 0 | 1 |
| RARS2 # | 0 | 1 |
| SCN1A* | 0 | 1 |
| SLC16A2 | 0 | 1 |
| STXBP1* | 0 | 1 |

**Supplementary Table 2: CVI-associated genetic diagnoses in 100KGP**

Key

* Genes already known to be associated with CVI on HPO website (https://hpo.jax.org/app/).

\# Genes represented in both the DECIPHER and 100KGP cohorts

| **Gene Name** | **Number of participants with CVI with a TIER 1 variant reported** | **Number of participants with CVI with a TIER 2 variant reported** |
| --- | --- | --- |
| PITRM1 | 2 | 1 |
| KCNT1 | 2 | 0 |
| CACNA1A | 1 | 3 |
| SHANK3* | 1 | 3 |
| KANSL1 | 1 | 2 |
| GRIN2B*# | 1 | 1 |
| WDR73 | 1 | 1 |
| TCF4*# | 1 | 0 |
| TUBB4A | 1 | 0 |
| TUBB2A | 1 | 0 |
| SRD5A3 | 1 | 0 |
| HNRNPU | 1 | 0 |
| NR2F1* | 1 | 0 |
| POLR3A | 1 | 0 |
| CSTB | 1 | 0 |
| SIN3A | 1 | 0 |
| GNAS | 1 | 0 |
| HECW2* | 1 | 0 |
| CBL | 1 | 0 |
| MTOR | 1 | 0 |
| SETD2 | 1 | 0 |
| SCN2A | 1 | 0 |
| UBA5 | 1 | 0 |
| DMD | 0 | 5 |
| MT-ATP6 | 0 | 5 |
| HIVEP2* | 0 | 3 |
| TRIO | 0 | 3 |
| CCDC22 | 0 | 3 |
| MT-ATP8 | 0 | 3 |
| KMT2D | 0 | 3 |
| FLNA | 0 | 3 |
| MT-ND5 | 0 | 2 |
| KAT6A*# | 0 | 2 |
| ANKRD11 | 0 | 2 |
| KMT2C | 0 | 2 |
| ZIC2* | 0 | 2 |
| KCNQ5* | 0 | 2 |
| CC2D2A* | 0 | 2 |
| COL4A2 | 0 | 2 |
| MT-CO1 | 0 | 2 |
| NIPBL | 0 | 2 |
| OFD1 | 0 | 2 |
| C5orf42 | 0 | 2 |
| NF1 | 0 | 2 |
| DHTKD1 | 0 | 2 |
| MT-CYB | 0 | 1 |
| SLC35A2* | 0 | 1 |
| MCCC1 | 0 | 1 |
| EIF2B5 | 0 | 1 |
| CHD7 | 0 | 1 |
| HCCS | 0 | 1 |
| DDX3X | 0 | 1 |
| FLVCR1 | 0 | 1 |
| ZNF711 | 0 | 1 |
| ATP13A2 | 0 | 1 |
| SZT2 | 0 | 1 |
| ABCB7 | 0 | 1 |
| ACSL4 | 0 | 1 |
| COQ4 | 0 | 1 |
| CHD4 | 0 | 1 |
| RERE* | 0 | 1 |
| ARID1A | 0 | 1 |
| AUTS2 | 0 | 1 |
| PHF8 | 0 | 1 |
| KNL1 | 0 | 1 |
| CHD8 | 0 | 1 |
| SMAD4 | 0 | 1 |
| BAG3 | 0 | 1 |
| HEPACAM | 0 | 1 |
| TTN | 0 | 1 |
| ALG13 | 0 | 1 |
| EBP | 0 | 1 |
| IFIH1 | 0 | 1 |
| RNF170 | 0 | 1 |
| ATP7B | 0 | 1 |
| SHANK2 | 0 | 1 |
| COL11A2 | 0 | 1 |
| ACTL6A | 0 | 1 |
| MT-ND1 | 0 | 1 |
| BSCL2 | 0 | 1 |
| OPHN1 | 0 | 1 |
| GK | 0 | 1 |
| ASXL3 | 0 | 1 |
| SPG7 | 0 | 1 |
| ATM | 0 | 1 |
| NDUFA1 | 0 | 1 |
| PUF60 | 0 | 1 |
| AFF2 | 0 | 1 |
| MTHFR | 0 | 1 |
| ERCC5 | 0 | 1 |
| MECP2 | 0 | 1 |
| IL1RAPL1 | 0 | 1 |
| TREX1 | 0 | 1 |
| ACY1 | 0 | 1 |
| SLC9A6 | 0 | 1 |
| DVL1 | 0 | 1 |
| DHX30 | 0 | 1 |
| SPTAN1 | 0 | 1 |
| PDHA1 # | 0 | 1 |
| AMER1 | 0 | 1 |
| UNC80 | 0 | 1 |
| FGD1 | 0 | 1 |
| MFN2 | 0 | 1 |
| DGUOK | 0 | 1 |
| SCAPER | 0 | 1 |
| KIF1A*# | 0 | 1 |
| TSEN2* | 0 | 1 |
| OTC | 0 | 1 |
| MCM3AP | 0 | 1 |
| ATP6AP1 | 0 | 1 |
| IKBKG | 0 | 1 |
| SYN1 | 0 | 1 |
| PIGA* | 0 | 1 |
| MYT1L* | 0 | 1 |
| CHAMP1* | 0 | 1 |
| TRIP12 | 0 | 1 |
| FOXG1 *# | 0 | 1 |
| KDM6A | 0 | 1 |
| CACNA1G | 0 | 1 |
| SMARCA2 | 0 | 1 |
| FBXO11 | 0 | 1 |
| PACS1 | 0 | 1 |
| MED13L | 0 | 1 |
| AFF4 | 0 | 1 |
| KCNK9 | 0 | 1 |
| NPRL3 | 0 | 1 |
| WDR45 | 0 | 1 |
| ROR2 | 0 | 1 |
| CHRNE | 0 | 1 |
| KIF11 | 0 | 1 |
| UBE3A | 0 | 1 |
| SMARCC2 | 0 | 1 |
| HDAC4 | 0 | 1 |
| FAT4 | 0 | 1 |
| SACS | 0 | 1 |
| FTCD | 0 | 1 |
| AFG3L2 | 0 | 1 |
| PTEN | 0 | 1 |
| RARS2 | 0 | 1 |
| RLIM | 0 | 1 |
| TSC2 | 0 | 1 |
| WFS1 | 0 | 1 |
| ZC4H2 | 0 | 1 |
| ARID1B | 0 | 1 |

**Supplementary Table 3: Literature review for CVI-associated genetic diagnoses present in both 100KGP and DECIPHER patients**

| **Gene** | **Gene function** | **Phenotype summary** | **Case series reference** | **Total number of patients in paper** | **Any visual phenotypes?** | **CVI reported** | **Other ophthalmological features reported** |
| --- | --- | --- | --- | --- | --- | --- | --- |
| *FOXG1* | Transcription factor | Complex neurodevelopmental syndrome, with Rett-like features | [1] | 8 | yes | 8/8 - fatigability of and deficits in visual attention and engagement |  |
|  |  |  | [2] | 122 | yes | CVI 41% | Strabismus 64% |
|  |  |  | [3] | 45 | yes | No | Strabismus, poor eye contact, abnormal ocular pursuit  16 / 45 as initial concerns  38 / 42 on examination |
|  |  |  | [4] | 26 | yes | No | Strabismus, poor eye contact in 9/9 where information available |
| *PDHA1* | Pyruvate dehydrogenase deficiency | Mitochondrial dysfunction | [5] | 371 | yes | No | Optic atrophy 4%  Nystagmus 3%  Strabismus 1.6% |
| *RARS2* | Mitochondrial | Pontocerebellar hypoplasia type 6 | [6] | 53 | Yes | visual impairment unspecified 19% | nystagmus 4%, optic atrophy 4% |
| *GRIN2B* | NMDA receptor subunit | Developmental and epileptic encephalopathy, with structural brain malformations | [7] | 48 new, 43 from lit | Yes | CVI 7% (a/w cortical malformations) |  |
| *TCF4* | Transcription factor | Pitt Hopkins Syndrome | [8] | 112 | yes | No | Strabismus 62%  Myopia 48% |
|  |  |  | [9] | 16 | yes | No | Strabismus  Myopia  Astigmatism |
|  |  |  | [10] | 10 | yes | No | Nystagmus, astigmatism, strabismus, myopia |
|  |  |  | [11] | 23 | yes | No | Myopia  strabismus |
|  |  |  | [12] | 100 | yes | No | Up to 50%  Strabismus  Myopia  Astigmatism |
| *KAT6A* | histone acetyltransferase, transcriptional regulation | Intellectual disability, Arboleda-Tham syndrome | [13] | 5 new, 80 from lit | yes | Visual defects 65% | Strabismus 57% |
|  |  |  | [14] | 76 | Yes | Visual defect 63% | Strabismus 54% |
| *KIF1A* | Kinesin motor protein, synaptic vesicle transport | Neurodegeneration and spasticity with or without cerebellar atrophy | [15] | 10 new, 99 literature cases | Yes | CVI in 3 / 10 new cases | Optic atrophy common |
|  |  |  | [16] | 28 new cases | yes |  | Optic atrophy 25% |

1. Boggio EM, Pancrazi L, Gennaro M, et al. Visual impairment in FOXG1-mutated individuals and mice. Neuroscience 2016;**324**:496-508 doi: 10.1016/j.neuroscience.2016.03.027[published Online First: Epub Date]|.

2. Brimble E, Reyes KG, Kuhathaas K, et al. Expanding genotype-phenotype correlations in FOXG1 syndrome: results from a patient registry. Orphanet J Rare Dis 2023;**18**(1):149 doi: 10.1186/s13023-023-02745-y[published Online First: Epub Date]|.

3. Vegas N, Cavallin M, Maillard C, et al. Delineating FOXG1 syndrome: From congenital microcephaly to hyperkinetic encephalopathy. Neurol Genet 2018;**4**(6):e281 doi: 10.1212/NXG.0000000000000281[published Online First: Epub Date]|.

4. Kortum F, Das S, Flindt M, et al. The core FOXG1 syndrome phenotype consists of postnatal microcephaly, severe mental retardation, absent language, dyskinesia, and corpus callosum hypogenesis. J Med Genet 2011;**48**(6):396-406 doi: 10.1136/jmg.2010.087528[published Online First: Epub Date]|.

5. Patel KP, O'Brien TW, Subramony SH, et al. The spectrum of pyruvate dehydrogenase complex deficiency: clinical, biochemical and genetic features in 371 patients. Mol Genet Metab 2012;**106**(3):385-94 doi: 10.1016/j.ymgme.2012.03.017[published Online First: Epub Date]|.

6. Zhang Y, Yu Y, Zhao X, et al. Novel RARS2 Variants: Updating the Diagnosis and Pathogenesis of Pontocerebellar Hypoplasia Type 6. Pediatr Neurol 2022;**131**:30-41 doi: 10.1016/j.pediatrneurol.2022.04.002[published Online First: Epub Date]|.

7. Platzer K, Yuan H, Schutz H, et al. GRIN2B encephalopathy: novel findings on phenotype, variant clustering, functional consequences and treatment aspects. J Med Genet 2017;**54**(7):460-70 doi: 10.1136/jmedgenet-2016-104509[published Online First: Epub Date]|.

8. Whalen S, Heron D, Gaillon T, et al. Novel comprehensive diagnostic strategy in Pitt-Hopkins syndrome: clinical score and further delineation of the TCF4 mutational spectrum. Hum Mutat 2012;**33**(1):64-72 doi: 10.1002/humu.21639[published Online First: Epub Date]|.

9. Marangi G, Ricciardi S, Orteschi D, et al. The Pitt-Hopkins syndrome: report of 16 new patients and clinical diagnostic criteria. Am J Med Genet A 2011;**155A**(7):1536-45 doi: 10.1002/ajmg.a.34070[published Online First: Epub Date]|.

10. Van Balkom ID, Vuijk PJ, Franssens M, et al. Development, cognition, and behaviour in Pitt-Hopkins syndrome. Dev Med Child Neurol 2012;**54**(10):925-31 doi: 10.1111/j.1469-8749.2012.04339.x[published Online First: Epub Date]|.

11. Goodspeed K, Newsom C, Morris MA, et al. Pitt-Hopkins Syndrome: A Review of Current Literature, Clinical Approach, and 23-Patient Case Series. J Child Neurol 2018;**33**(3):233-44 doi: 10.1177/0883073817750490[published Online First: Epub Date]|.

12. Zollino M, Zweier C, Van Balkom ID, et al. Diagnosis and management in Pitt-Hopkins syndrome: First international consensus statement. Clin Genet 2019;**95**(4):462-78 doi: 10.1111/cge.13506[published Online First: Epub Date]|.

13. Urreizti R, Lopez-Martin E, Martinez-Monseny A, et al. Five new cases of syndromic intellectual disability due to KAT6A mutations: widening the molecular and clinical spectrum. Orphanet J Rare Dis 2020;**15**(1):44 doi: 10.1186/s13023-020-1317-9[published Online First: Epub Date]|.

14. Kennedy J, Goudie D, Blair E, et al. KAT6A Syndrome: genotype-phenotype correlation in 76 patients with pathogenic KAT6A variants. Genet Med 2019;**21**(4):850-60 doi: 10.1038/s41436-018-0259-2[published Online First: Epub Date]|.

15. Montenegro-Garreaud X, Hansen AW, Khayat MM, et al. Phenotypic expansion in KIF1A-related dominant disorders: A description of novel variants and review of published cases. Hum Mutat 2020;**41**(12):2094-104 doi: 10.1002/humu.24118[published Online First: Epub Date]|.

16. Vecchia SD, Tessa A, Dosi C, et al. Monoallelic KIF1A-related disorders: a multicenter cross sectional study and systematic literature review. J Neurol 2022;**269**(1):437-50 doi: 10.1007/s00415-021-10792-3[published Online First: Epub Date]|.

**Supplementary Table 4: Top 20 HPO terms in DECIPHER CVI cohort and control group.**

Bold HPO terms= significant after correction for multiple comparisons

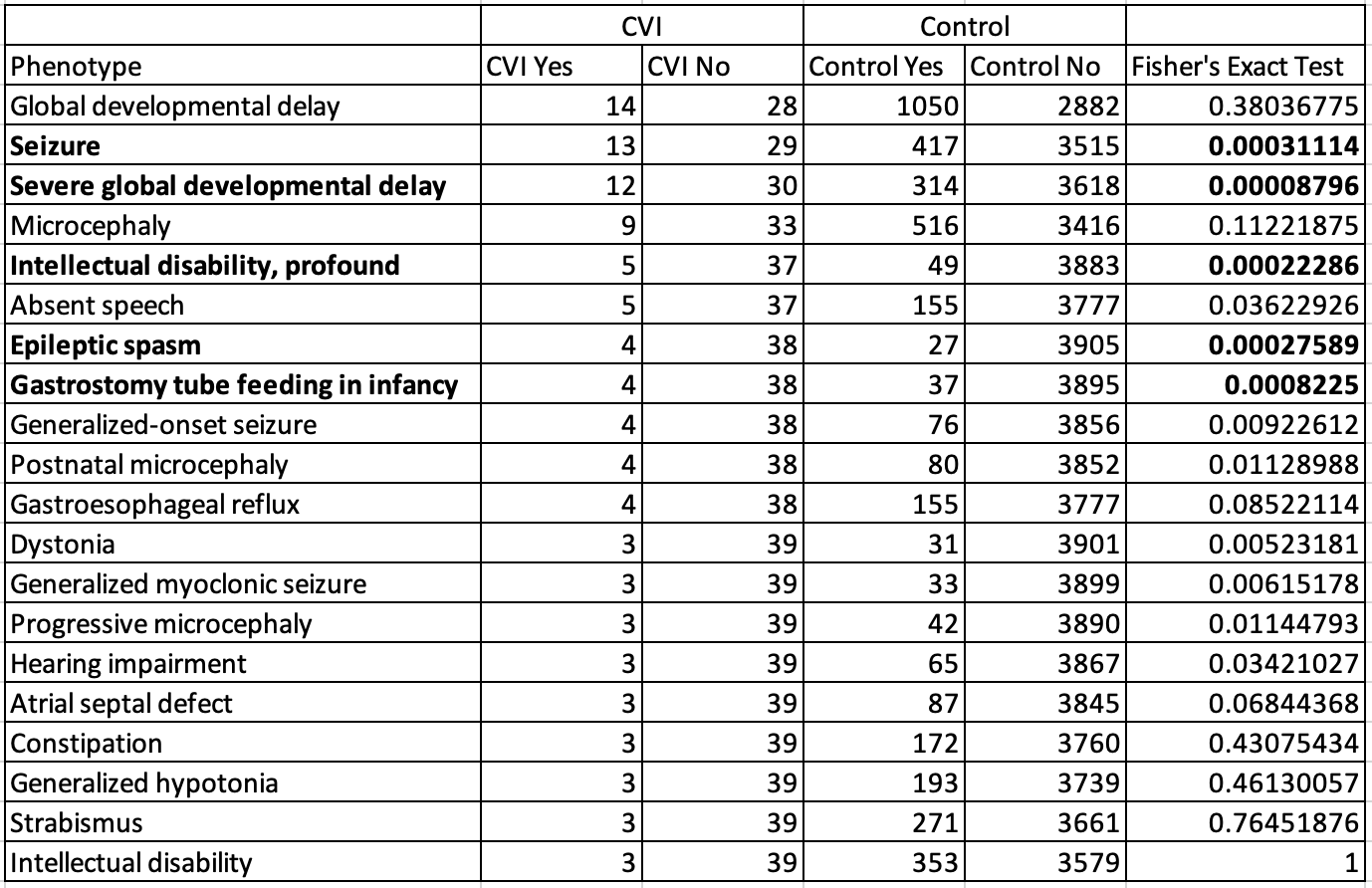

**Supplementary Table 5: Top 20 HPO terms in 100KGP CVI and control groups.**

Bold HPO terms= significant after correction for multiple comparisons

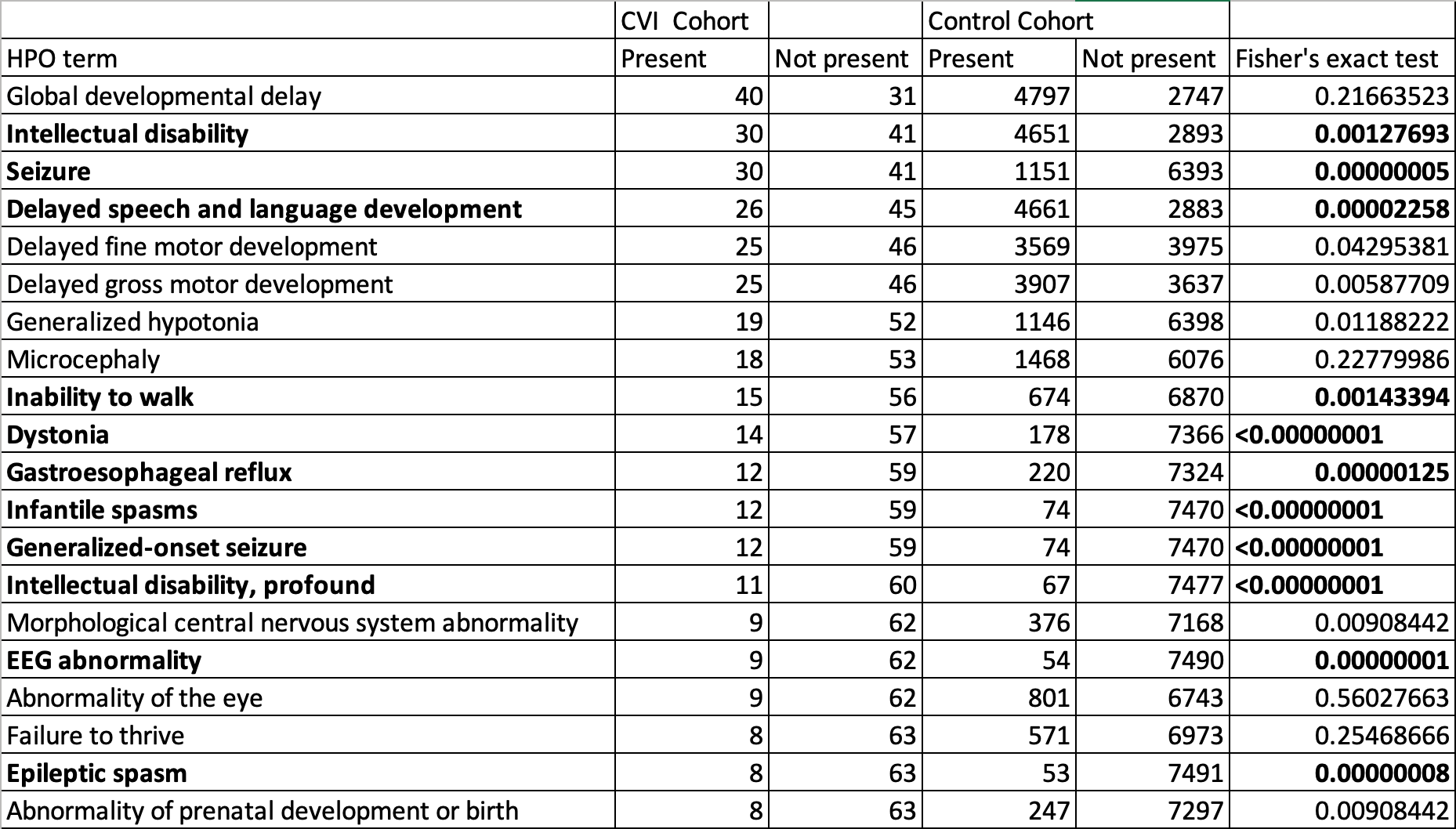

**Supplementary Figure 1: ShinyGO enrichment networks of control gene sets in DECIPHER**

(6 gene sets did not produce networks)

| 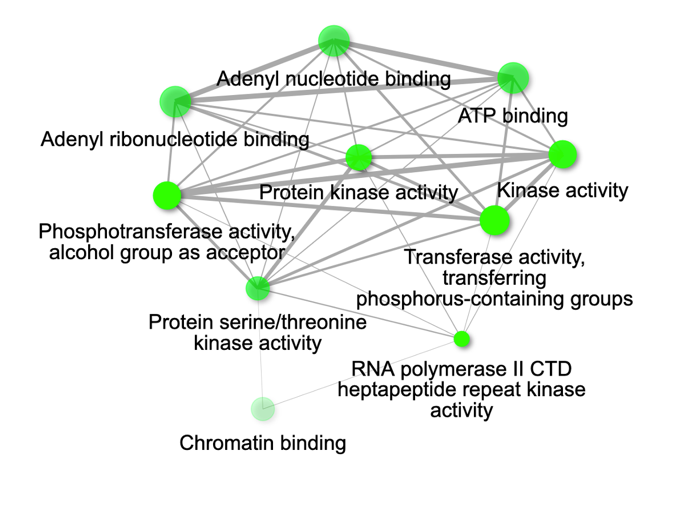 | 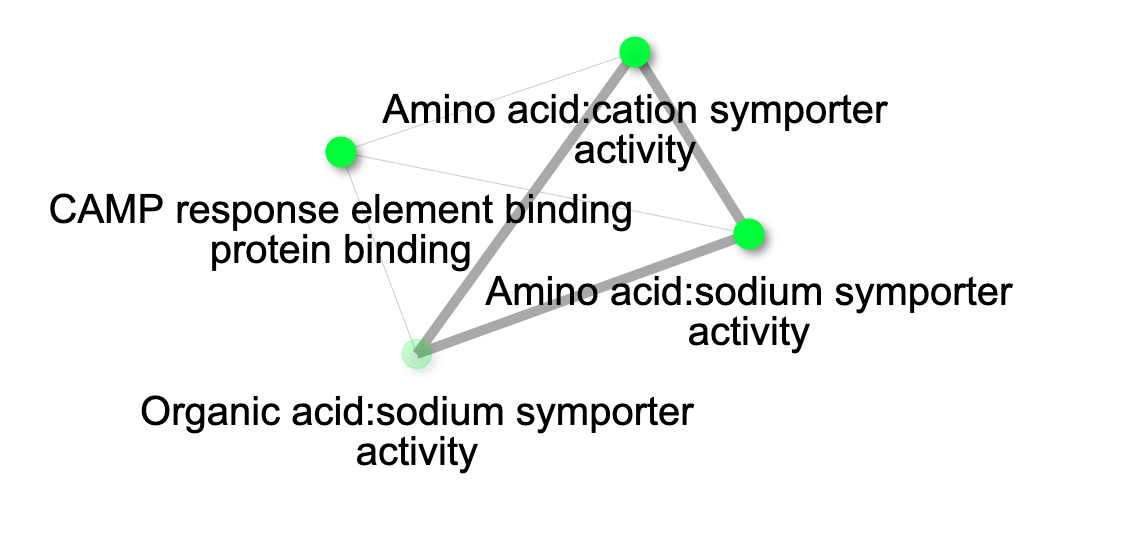 |
| --- | --- |
| 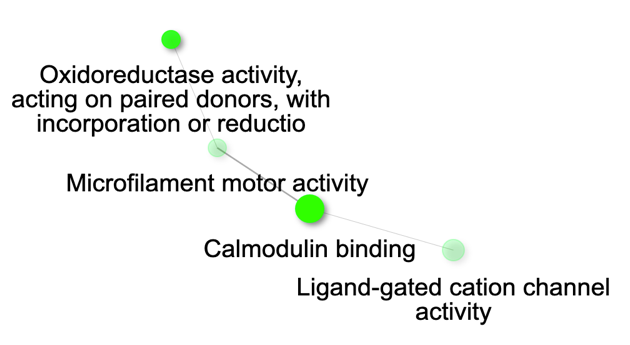 | 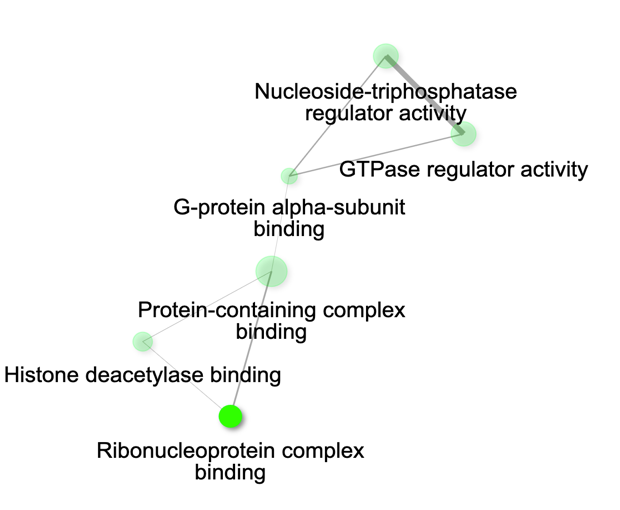 |

**Supplementary Figure 2: ShinyGO enrichment networks of control gene sets in 100KGP**

| 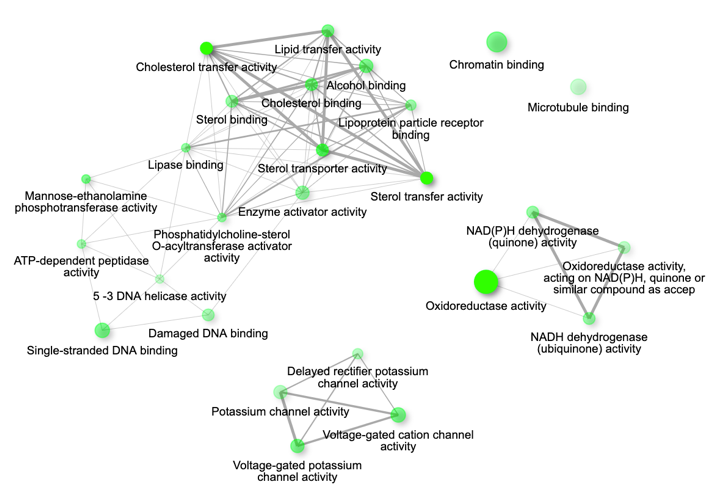 | 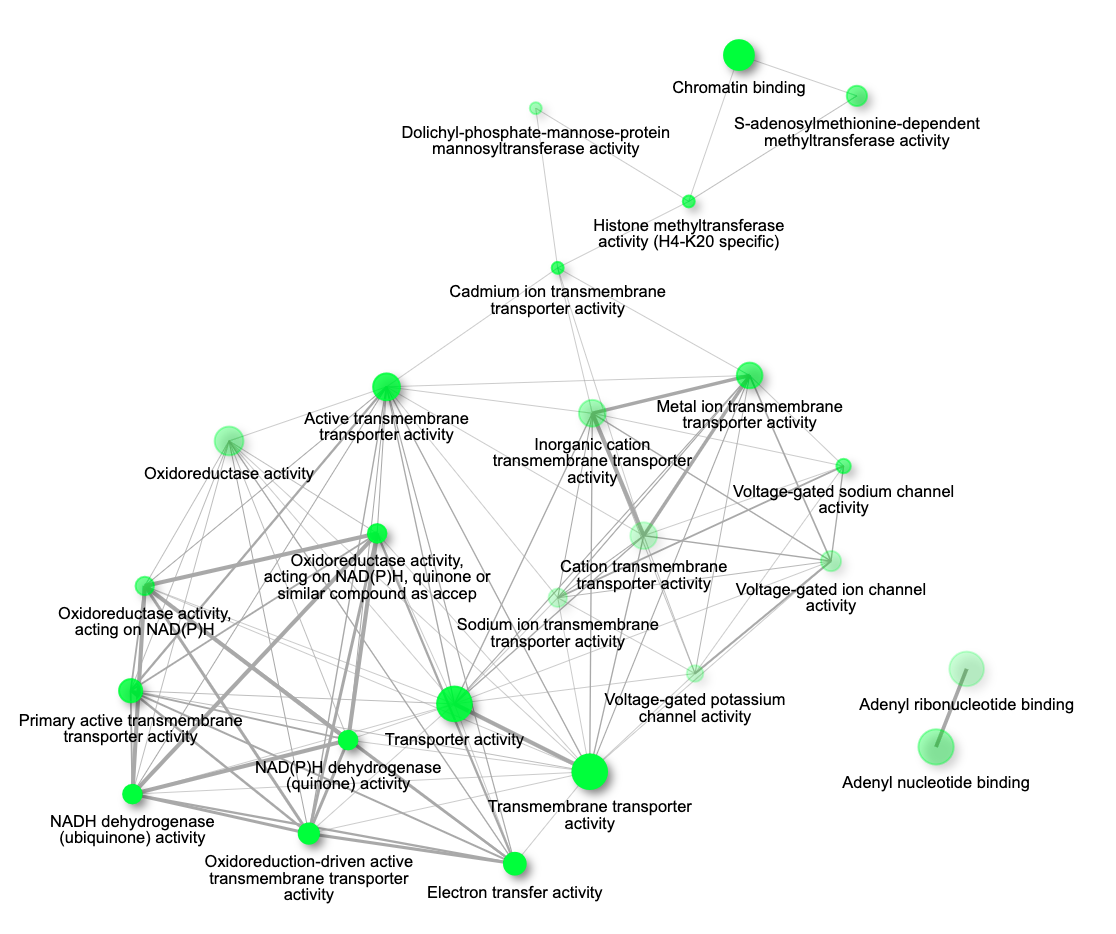 |
| --- | --- |
| 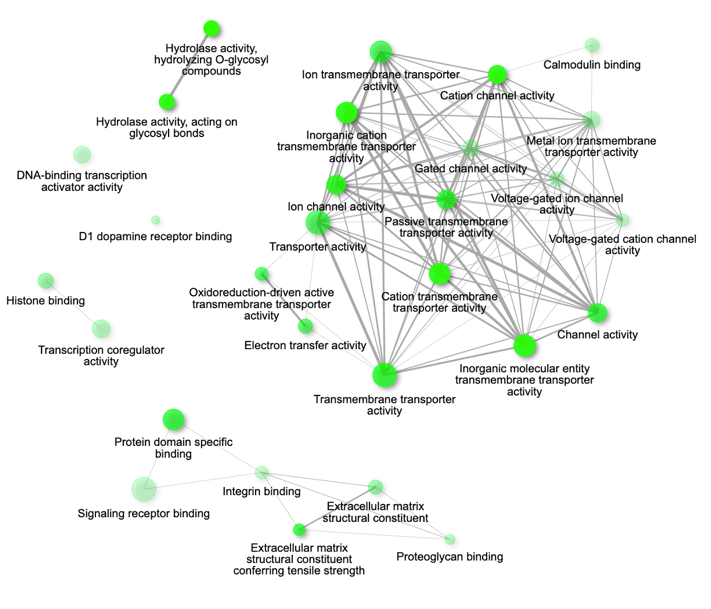 | 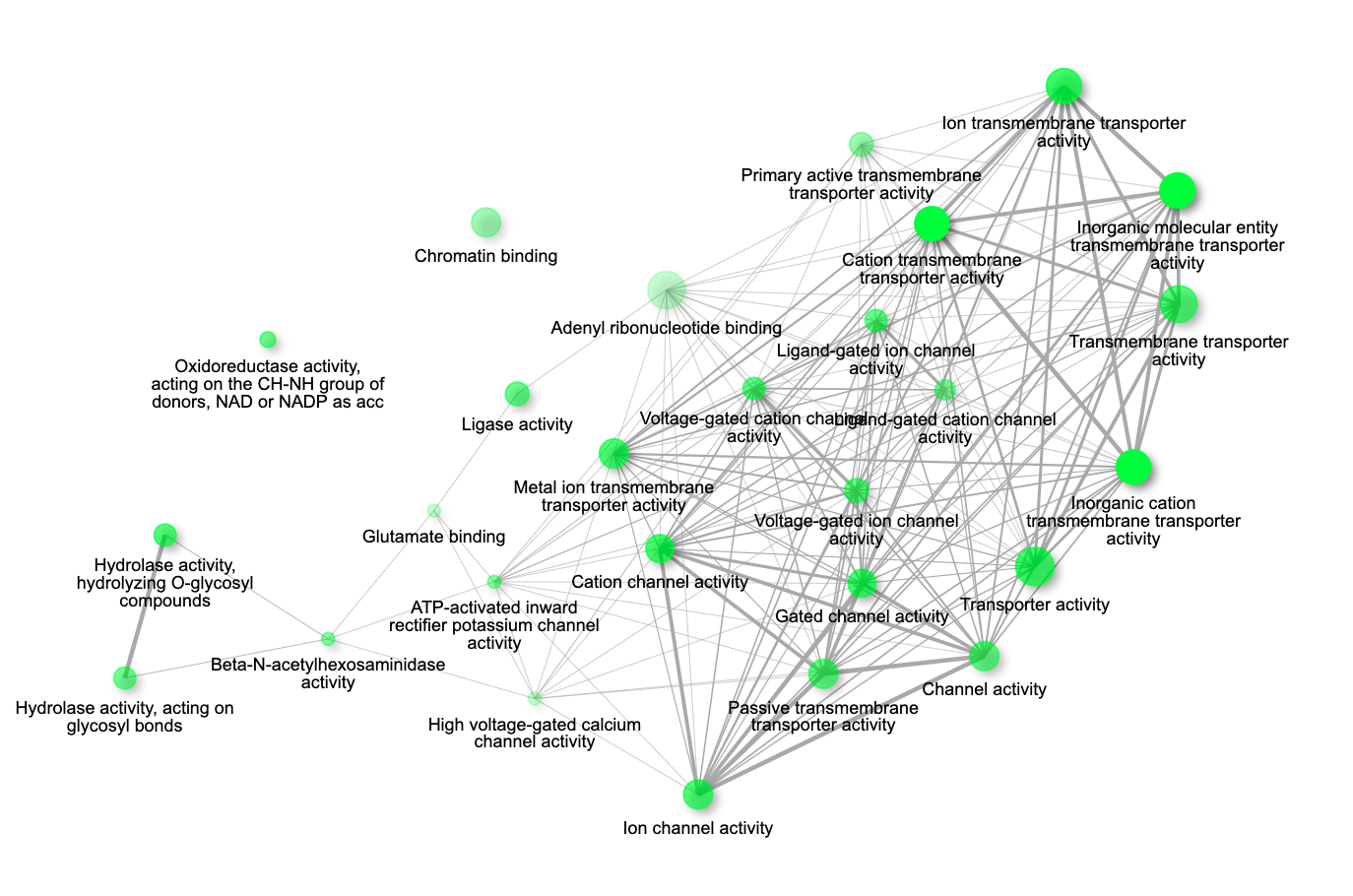 |
| 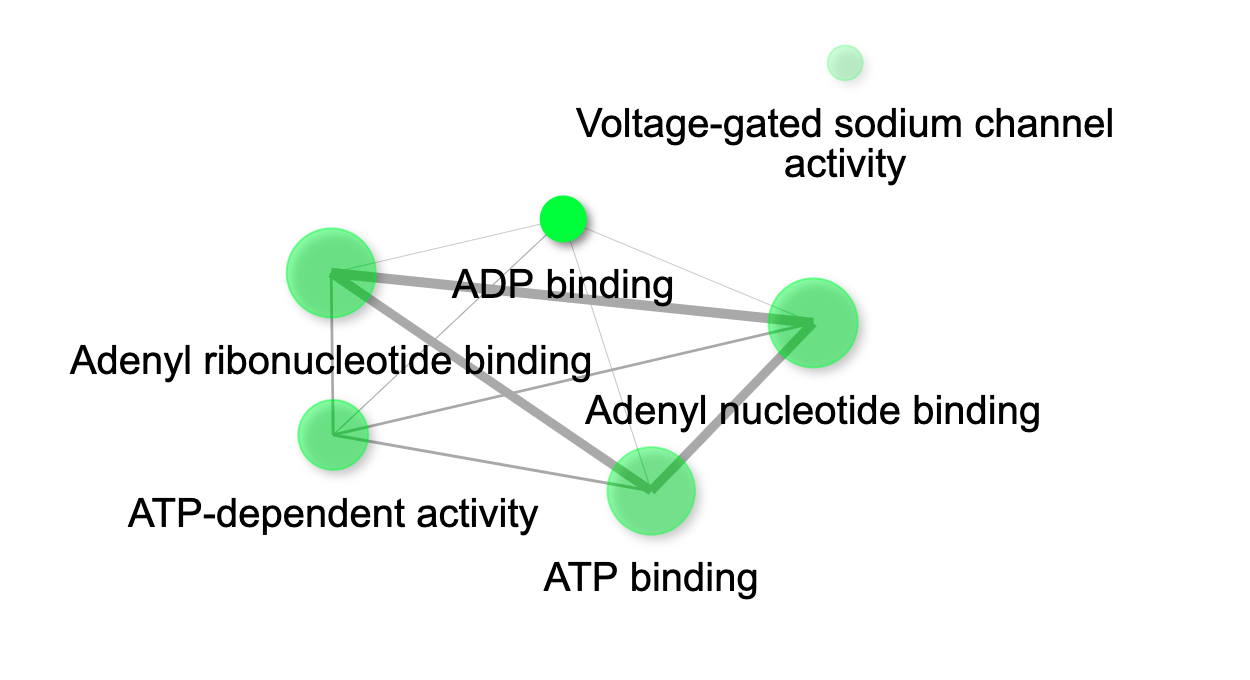 | 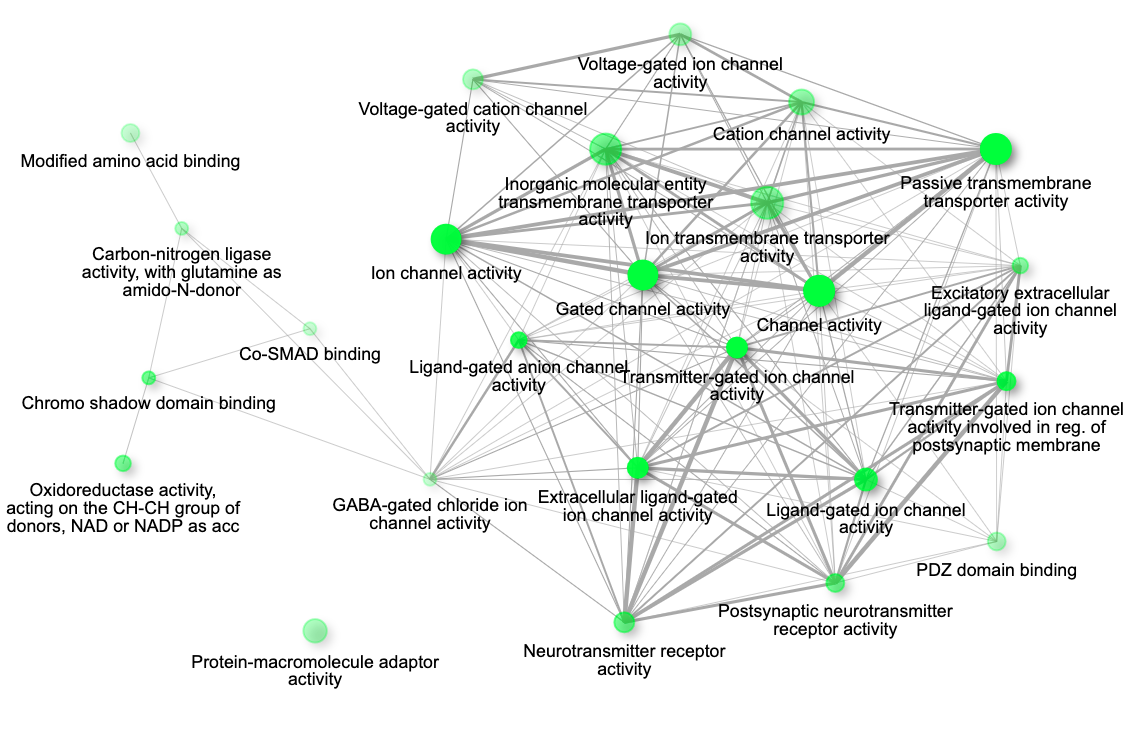 |
| 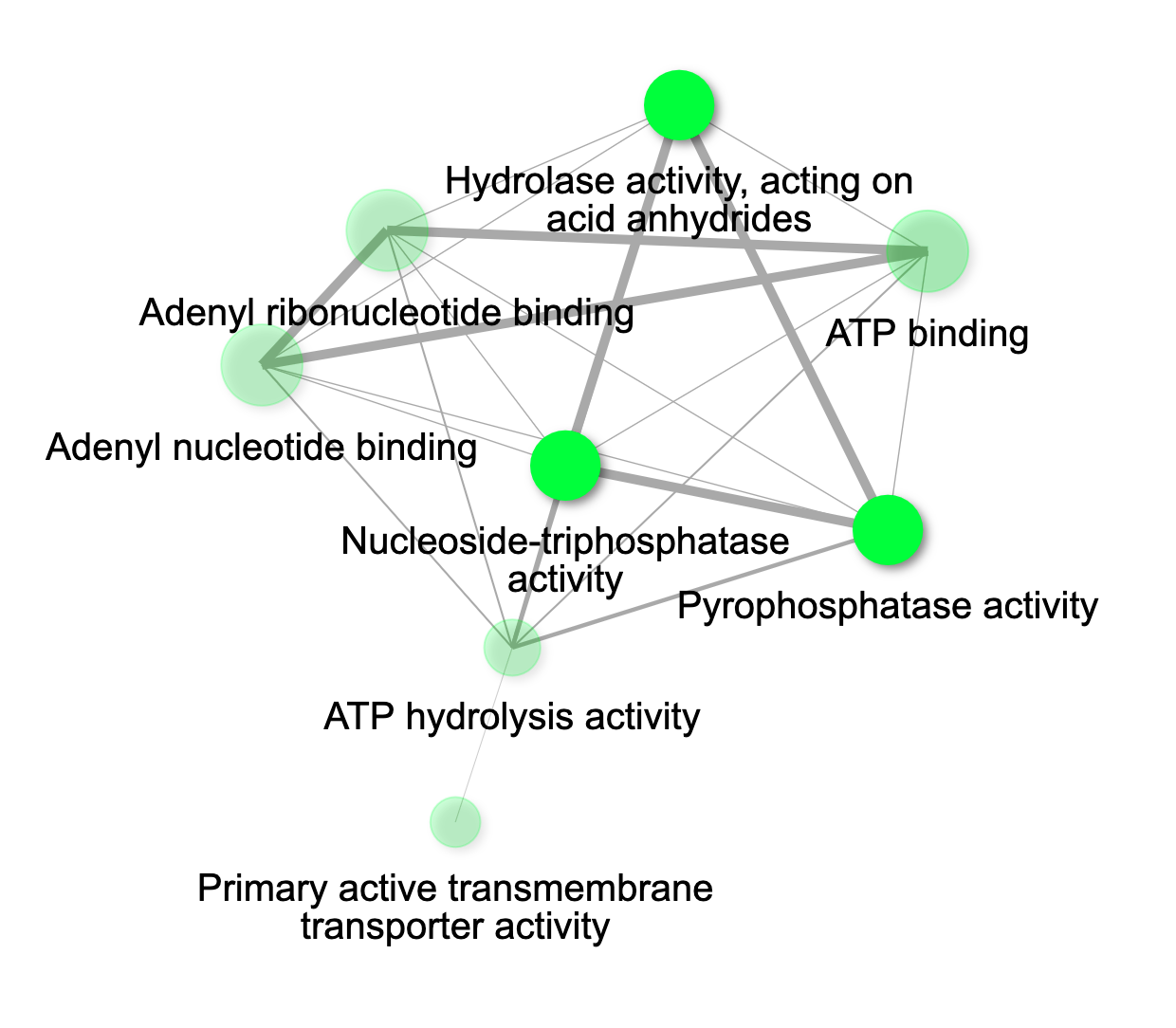 | 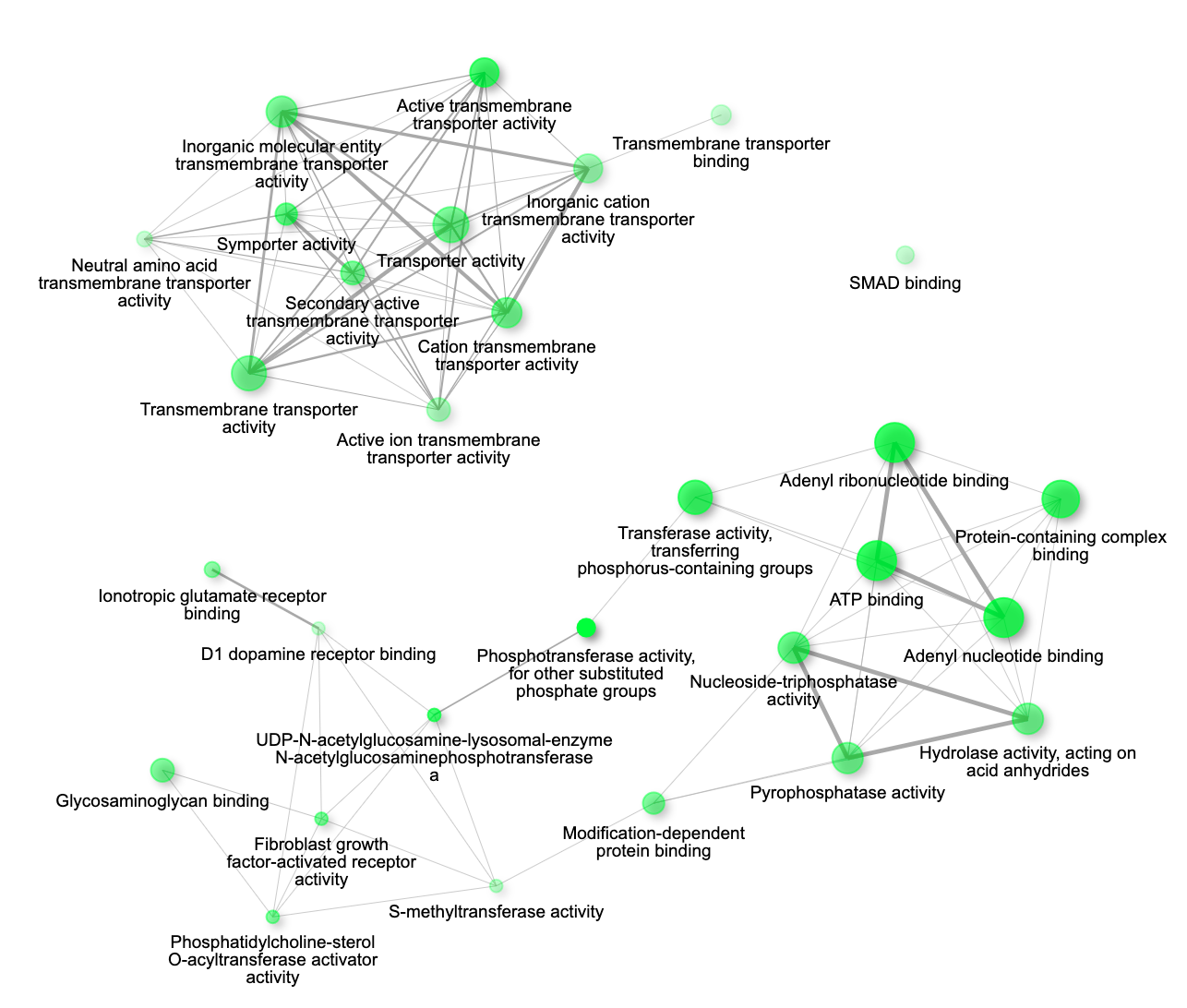 |
| 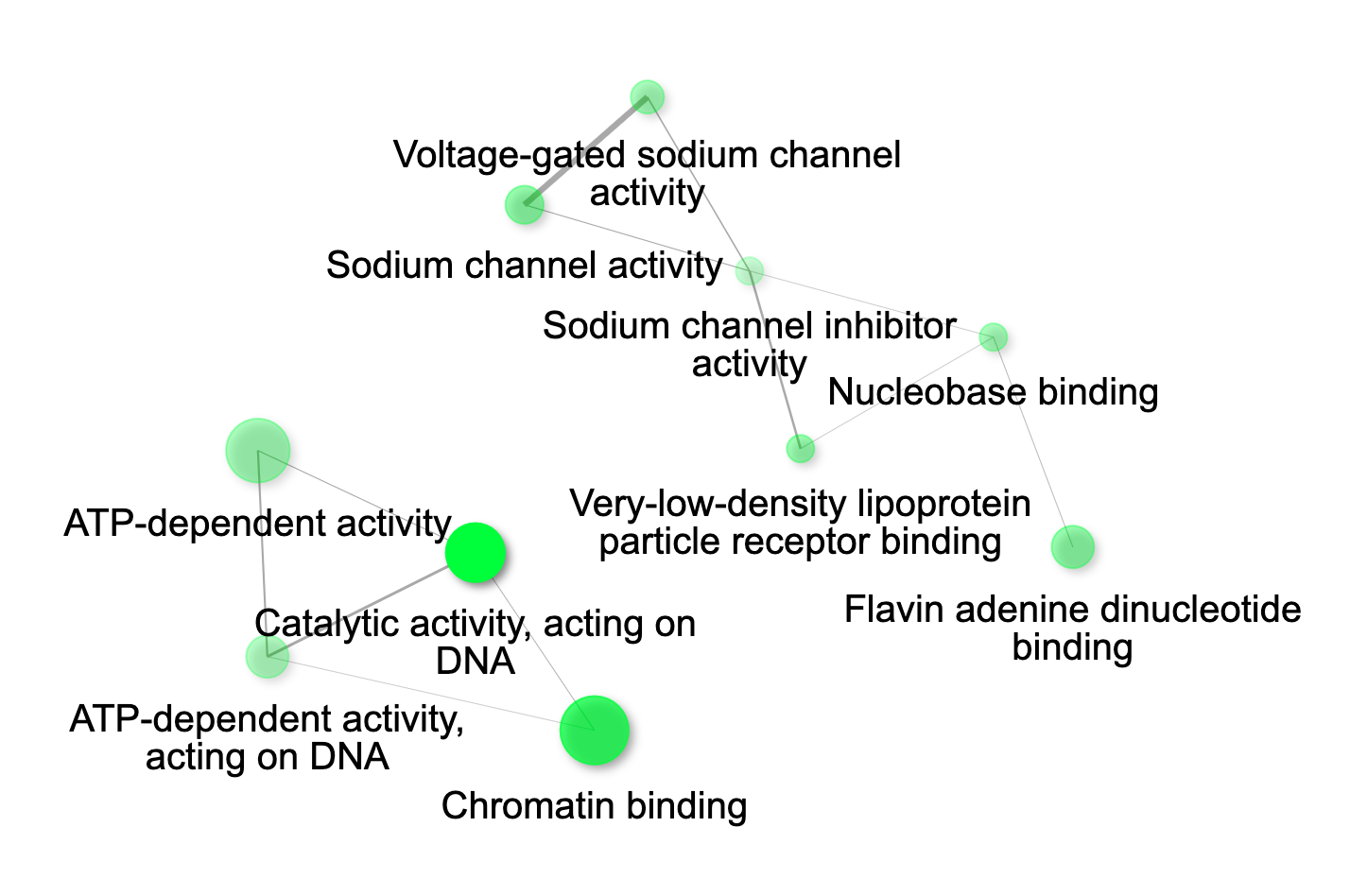 | 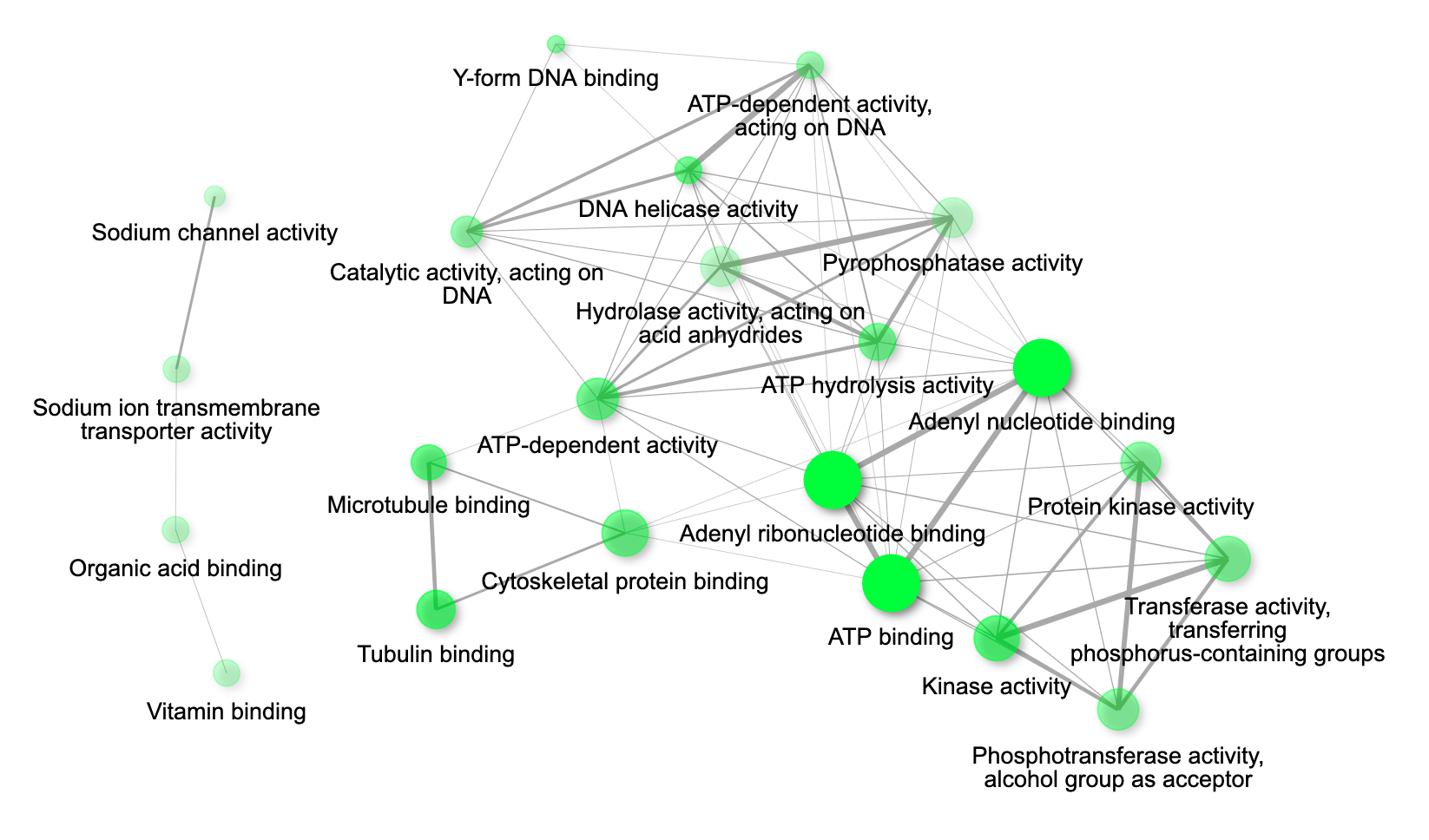 |
